# Peer-assisted HIV partner notification services to strengthen index partner testing for newly diagnosed men who have sex with men in coastal Kenya

**DOI:** 10.1101/2025.05.16.25327545

**Authors:** Haley Adrian, Maartje Dijkstra, Shally Mahmoud, Khamisi Mohamed, Evans Gichuru, Eduard J. Sanders, Don Operario, Elise M. van der Elst

## Abstract

**Background:** HIV partner notification services (PNS) have been proven safe and effective in finding undiagnosed HIV infections among general populations in settings with high HIV incidence, but have not been regularly implemented among men who have sex with men in sub-Saharan Africa. This study aimed to describe experiences with PNS in a cohort of newly diagnosed MSM in coastal Kenya, assess facilitators and barriers to participating in PNS, and explore which PNS strategies were preferred for different types of sexual partners.

**Methods:** The mixed-method study was conducted between January and July 2019 involving 27 MSM participants newly diagnosed with HIV who participated in PNS, of which 18 accepted to participate in a semi-structured in-depth interview which captured their perceptions, experiences, and views on PNS. Inductive thematic analysis was used to analyze qualitative data.

**Results:** The median age of participants was 28 (interquartile range [IQR] 25-36) years old, and 44.4% completed primary school. The median number of named sexual partners in the previous 12 months was 3 (IQR 2-6; total partners 109). Facilitators to participation in PNS included reassurance of personal safety, support from peer-mobilizers, and a sense of responsibility to others’ well-being to prevent HIV transmission. Barriers to PNS participation included fear of stigma and discrimination as well as missing or incorrect partner contact information. Provider-assisted partner notification was the preferred strategy selected by participants across all types of sexual partners. No participant reported experiencing any IPV or other social harms.

**Conclusions:** These findings suggest that PNS, particularly provider-assisted PNS, is a safe and promising HIV testing and linkage strategy for use with MSM in coastal Kenya.

## Introduction

An important gap in HIV prevention is the large proportion of persons living with HIV (PLHIV) who are unaware of their infection [1]. In order to reach the UNAIDS 95-95-95 global HIV targets - particularly the target that 95% of all PLHIV know their HIV status - innovative approaches are needed to increase HIV testing and linkage in unreached, high-incidence populations, such as men who have sex with men (MSM) [2].

Partner notification services (PNS) have been proven to be a safe and effective strategy for identifying people with undiagnosed HIV among the general population in several high HIV-incidence countries in Sub-Saharan Africa (SSA) [3, 4, 5]. PNS is a process by which the sexual partner(s) of an index client (i.e., an individual newly diagnosed with HIV) are notified of their risk of exposure to HIV and recommended to seek HIV testing. In addition to facilitating HIV testing, PNS also provides access to preventative HIV-related services (i.e., PrEP) or treatment for HIV (i.e., ART), as needed. PNS is a voluntary process that initiates once consent from the index client is received and is typically delivered by trained health care providers or trained lay persons (i.e., peer mobilizers). Several strategies are used for partner notification, which include both passive and assisted approaches: 1) Index referral: the index client notifies his or her sexual partner(s) and recommends HIV testing or provides the partner(s) with an HIV oral self-test, with optional disclosure of HIV status by the index client; 2) Provider referral or assisted partner notification services: the index client discloses contact information of his or her sexual partner(s) to a health provider who then attempts to make contact with the sexual partner(s) and offer voluntary HIV testing. Optimally, the health provider and index client consider the potential risk of intimate partner violence (IPV) and other social harms before deciding whether to continue with selecting a PNS strategy. If determined to have little to no risk of social harms, the health provider then contacts the partner(s), informs them of their risk of exposure for HIV and recommends HIV testing. This can be done either with or without disclosure of the index client, depending on the preference of the index client [6].

Provider referral is the most effective PNS strategy without posing a risk to the index client and the strategy recommended by the WHO [7]. However, at the time of this study, no data was available on the effectiveness of different PNS strategies among MSM in SSA Africa, which is why this study allowed index clients to choose and explore all potential PNS strategies.

In high-resource settings, targeted PNS for MSM newly diagnosed with HIV has been shown to increase detection of undiagnosed HIV infections and enable initiation of ART and prevention services among MSM [8–14]. However, data on implementation, safety, and feasibility of offering PNS among MSM resource-limited settings such as sub-Saharan Africa are limited [15–19].

In Kenya, it is now national policy to offer assisted PNS to anyone who has tested positive for HIV [20]. Despite the existence of this national policy, PNS has solely been studied and proven effective and safe among heterosexual populations in Kenya. While PNS has the potential to benefit MSM in Kenya when conducted ethically (i.e., with consent and without coercion), it also has the potential to be implemented in ways that can cause social and physical harm to individuals, undermine their rights to consent, privacy, safety, and confidentiality, and erode trust with health care providers [21]. Due to experiences of stigma and discrimination in healthcare settings, many MSM in Kenya actively avoid engaging in HIV testing and other preventative health services [22]. Healthcare avoidance by MSM has been observed elsewhere in SSA [23, 24].

In this study, we set out to explore experiences with PNS and assess facilitators and barriers to participating in PNS among a sample of MSM newly diagnosed with HIV in coastal Kenya, in order to improve uptake of PNS and ultimately increase HIV case-finding. Based on previous literature on the importance of peers in mobilizing and providing HIV prevention outreach to MSM populations [25, 26, 27], we hypothesized that peer-involved strategies would be identified as an essential factor facilitating the uptake of PNS among MSM [16]. This study was part of a larger study with newly-diagnosed MSM and their sexual partners in coastal Kenya [16], which found that PNS was a safe and effective method in identifying undiagnosed HIV infections. Here, we also wanted to explore whether index clients’ preferences for PNS strategies varied according to types of sexual partners, whether specific PNS strategies were more successful than others at enrolling sexual partners, and whether PNS was more successful at reaching specific types of sexual partners.

## Methods

We analyzed data from a mixed-methods study that aimed to evaluate the acceptability, feasibility, and safety of PNS among MSM newly diagnosed with HIV in coastal Kenya [12, 15]. Data were collected between April 3 – August 21, 2019. Briefly, we present descriptive quantitative data on sociodemographic variables and PNS characteristics for all index participants and their reported recent sexual partners (Tables 1 and 2). Next, we present thematic findings from qualitative analysis of individual interviews with index participants. All participants provided written informed consent prior to enrollment. The Kenya Medical Research Institute (KEMRI) Scientific Ethical Review Unit approved the study (135/3747).

### Study procedures

Research activities took place at the KEMRI clinics in Mtwapa and Malindi, coastal Kenya. The two sampling strategies used to engage participants were peer mobilization and PNS. MSM who were newly diagnosed with HIV at these sites were recruited in-person to the study by health care providers following receipt of their HIV diagnosis. Newly diagnosed HIV was defined as 2 positive rapid antibody results in a previous seronegative individual, or a positive point-of-care qualitative HIV RNA test (GeneXpert, Cepheid) in a seronegative or discordant individual (one test positive, one test negative) [16]. . These individuals were considered “index participants” in this study. After providing written informed consent, index participants completed an initial staff-administered interview survey during which participants were asked to disclose the total number of sexual partners in the previous 12 months and further details of each partner (i.e. type of partner [steady, casual, or sex work client] and gender identity of the partner). Using protocols developed previously, [28] staff performed IPV screening or facilitated index participants to self-assess for IPV in order to determine potential risks of harm for each partnership identified. If the risk of IPV and other harms was low to moderate, the index participant and staff discussed and determined the best strategy of partner notification (including index referral or provider referral – both strategies could include the provision of an oral self-test to the partner and the support of a peer mobilizer). For each partner, a different notification strategy could be chosen. For high-risk partnerships, no notification services were conducted. If the index participant opted for provider referral, index participants were asked to disclose names and contact information of their sexual partners. If partner contact details were unknown or missing, peer mobilizers and providers utilized other strategies (e.g., outreach at known hotspots) to attempt to reach the partner(s).

Approximately 6 weeks after the initial face-to-face interview with the index participant, a second structured face-to-face interview was conducted with the index participant to explore their experience with the PNS process using in-depth qualitative questions. The index participant was asked about his perceptions, experiences, and views on the partner notification services and whether he experienced IPV or other (social) harms.

In addition, sexual partners who were successfully contacted and notified were recruited into the study and additional demographic data of these partners were collected. Once enrolled, they were tested for HIV and classified as HIV negative, newly HIV diagnosed or known HIV positive. Sexual partners of index participants who were newly diagnosed with HIV could also enroll as an index participant and participate in this study.

### Operationalization of PNS Strategies

Protocols for this study guided the delivery of partner notification approaches. When the index participant opted for index referral, the provider and peer-mobilizers always supported and assisted the index participant throughout the process of notifying their partner(s), however, the notification was still done by the index. When the index participant opted for provider referral, the notification was done by the provider (with support of the peer mobilizer).

### Data management and analysis

Descriptive analyses were conducted to describe quantitative data for all 27 index participants enrolled, the total number of recent sexual partners they reported having and their partner characteristics.

Analysis of qualitative data utilized inductive thematic analysis, as described by Braun and Clarke [29]. This involved systematic coding, identifying, and defining concepts emerging from the data across the data set, mapping the concepts, creating typologies, finding associations between concepts, and seeking explanations from the data. Data were managed using NVivo (version 12). The primary researcher (H.A) independently coded the transcripts and created the codebook used for the analysis, which was then discussed, revised, and verified by two additional senior qualitative researchers (E.M.E, D.O) on the study. Any differences in coding were reconciled through group discussion with the larger research team (E.J.S., M.D). Team members reflected on their primary assumptions and positionalities in order to minimize interpretive bias (e.g., portraying PNS in a favorable light) and allow participants’ narratives to guide analysis and interpretations.

## Results

### Participant Characteristics

Overall, 27 index participants newly diagnosed with HIV consented to participate in PNS. Of the total 27 who enrolled, 18 participants (10 from Malindi, 8 from Mtwapa) further agreed to participate in the 2^nd^ phase of data collection involving semi-structured in-depth interviews to discuss their experiences with PNS.

Of the 27 total index participants who engaged in PNS, 55.6% identified as gay and 44.4% identified as bisexual (Table 1). The median age of participants was 28 (interquartile range [IQR] 25-36) years old, and the majority reported either primary (44.4%) or secondary level (44.4%) education. Most (70.4%) of the 27 participants were mobilized through peer-led outreach. There were no substantial differences between index participants who did (n=18) and did not (n=9) engage in the follow-up in-depth interview.

**Table 1.** Characteristics of Index Participants (n=27)

|  | Index Participant Category |  |  |  |  |  |
| --- | --- | --- | --- | --- | --- | --- |
|  | Did not participate in an IDI |  | Participated in an IDI |  | Total |  |
|  | (N=9) |  | (N=18) |  | (N=27) |  |
|  | N or median | % or IQR | N or median | % or IQR | N or median | % or IQR |
| Age (years) | 28 | 25-37 | 29 | 26-32 | 28 | 25-36 |
| Gender |  |  |  |  |  |  |
| Male | 9 | 100% | 18 | 100% | 28 | 100% |
| Sexuality |  |  |  |  |  |  |
| Gay | 6 | 66.7% | 9 | 50.0% | 15 | 55.6% |
| Bisexual | 3 | 33.3% | 9 | 50.0% | 12 | 44.4% |
| Education Level |  |  |  |  |  |  |
| Primary | 5 | 55.6% | 7 | 38.9% | 12 | 44.4% |
| Secondary | 4 | 44.4% | 8 | 44.4% | 12 | 44.4% |
| Vocational | 0 | 0% | 3 | 16.7% | 3 | 11.1% |
| Mobilization Strategy |  |  |  |  |  |  |
| Peer mobilization | 4 | 44.4% | 15 | 83.3% | 19 | 70.4% |
| PNS | 4 | 44.4% | 1 | 5.6% | 5 | 18.5% |
| Research clinic | 1 | 11.1% | 2 | 11.1% | 3 | 11.1% |
| Days between Initiation of PNS and IDI | - | - | 45 | 37-63 | - | - |
| Number of partners named | 3 | 3-4 | 4 | 2-9 | 3 | 2-6 |

The median number of sexual partners in the previous 12 months named by each of the participants was 3 (IQR 2-6), with a total number of 109 sexual partners named. Of these 109 partners, 56.9% were described by index participants as casual sexual partners, 22.9% as sex work clients (both regular and one-off clients), and 15.6% as steady partners (Table 2). Of the 109 named sexual partners, 65.1% were male, 19.3% were female, and 15.6% were transgender women. For the majority of partners (58.7%), the index participants chose to use provider-assisted PNS, compared to only 12.8% of partners for whom the index participant chose index referral. For over one-quarter (28.4%) of partners named, there was no notification strategy chosen for the following reasons: these partners were already enrolled in the study (n=19), contact details were unavailable (n=11), or the index participant was afraid of potential impacts (e.g., economic consequences) of disclosing this information (n=1).

**Table 2.** Characteristics of Recent Sexual Partners (N=109) identified by Index Participants (n=27)

|  | Steady<br>(N=17) |  | Casual<br>(N=62) |  | Client<br>(N=25) |  | Unknown<br>(N=5) |  | Total<br>(N=109) |  |
| --- | --- | --- | --- | --- | --- | --- | --- | --- | --- | --- |
|  | N | % | N | % | N | % | N | % | N | % |
| Partner Gender Identity |  |  |  |  |  |  |  |  |  |  |
| Male | 7 | 41.2% | 45 | 72.6% | 15 | 60.0% | 4 | 80.0% | 71 | 65.1% |
| Female | 9 | 52.9% | 8 | 12.9% | 4 | 16.0% | 0 | 0% | 21 | 19.3% |
| Transgender woman | 1 | 5.9% | 9 | 14.5% | 6 | 24.0% | 1 | 20.0% | 17 | 15.6% |
| PNS Strategy Chosen by Index Participant |  |  |  |  |  |  |  |  |  |  |
| Provider referral | 8 | 47.1% | 43 | 69.4% | 11 | 44.0% | 2 | 40.0% | 64 | 58.7% |
| Index referral | 5 | 29.4% | 5 | 8.1% | 3 | 12.0% | 1 | 20.0% | 14 | 12.8% |
| No PNS | 4 | 23.5% | 14 | 22.6% | 11 | 44.0% | 2 | 40.0% | 31 | 28.4% |
| Partner Enrolled | 5 | 29.4% | 27 | 43.6% | 9 | 36.0% | 3 | 60.0% | 44 | 40.4% |
| Age of Partner (years) | 24 | 23-31 | 27 | 24-23 | 29 | 27-44 | 26 | 21-30 | 27 | 24-32 |
| HIV Status of Partner |  |  |  |  |  |  |  |  |  |  |
| HIV negative | 1 | 20.0% | 10 | 37.1% | 1 | 11.1% | 3 | 100% | 15 | 34.1% |
| Newly HIV diagnosed | 0 | 0% | 7 | 25.9% | 3 | 33.3% | 0 | 0% | 10 | 22.7% |
| Know HIV positive | 4 | 80.0% | 10 | 37.1% | 5 | 55.6% | 0 | 0% | 19 | 43.2% |

Provider referral was the more commonly selected PNS strategy by index participants for reaching casual partners (69.4%) compared with steady partners (47.1%), whereas index referral was more commonly selected by index participants for reaching steady partners (29.4%) compared with casual partners (8.1%). Over half (56.4%) of partners who were notified were successfully enrolled in the study. Among those 44 sexual partners who were contacted through any PNS strategy and who enrolled in the study, 43.2% already knew of their HIV positive status, 34.1% tested negative for HIV, and 22.7% were newly diagnosed with HIV. Of the 10 sexual partners newly diagnosed with HIV, 70.0% were clients of index participants and 30.0% were casual sexual partners; no steady sexual partners were newly diagnosed.

### Emergent Themes from Qualitative Interviews

We identified five main domains describing barriers and facilitators to PNS based on follow-up in-depth interviews with PNS index participants, conducted 6 weeks after study enrollment. Table 3 outlines these domains and presents further sub-themes and representative quotes.

**Table 3.**
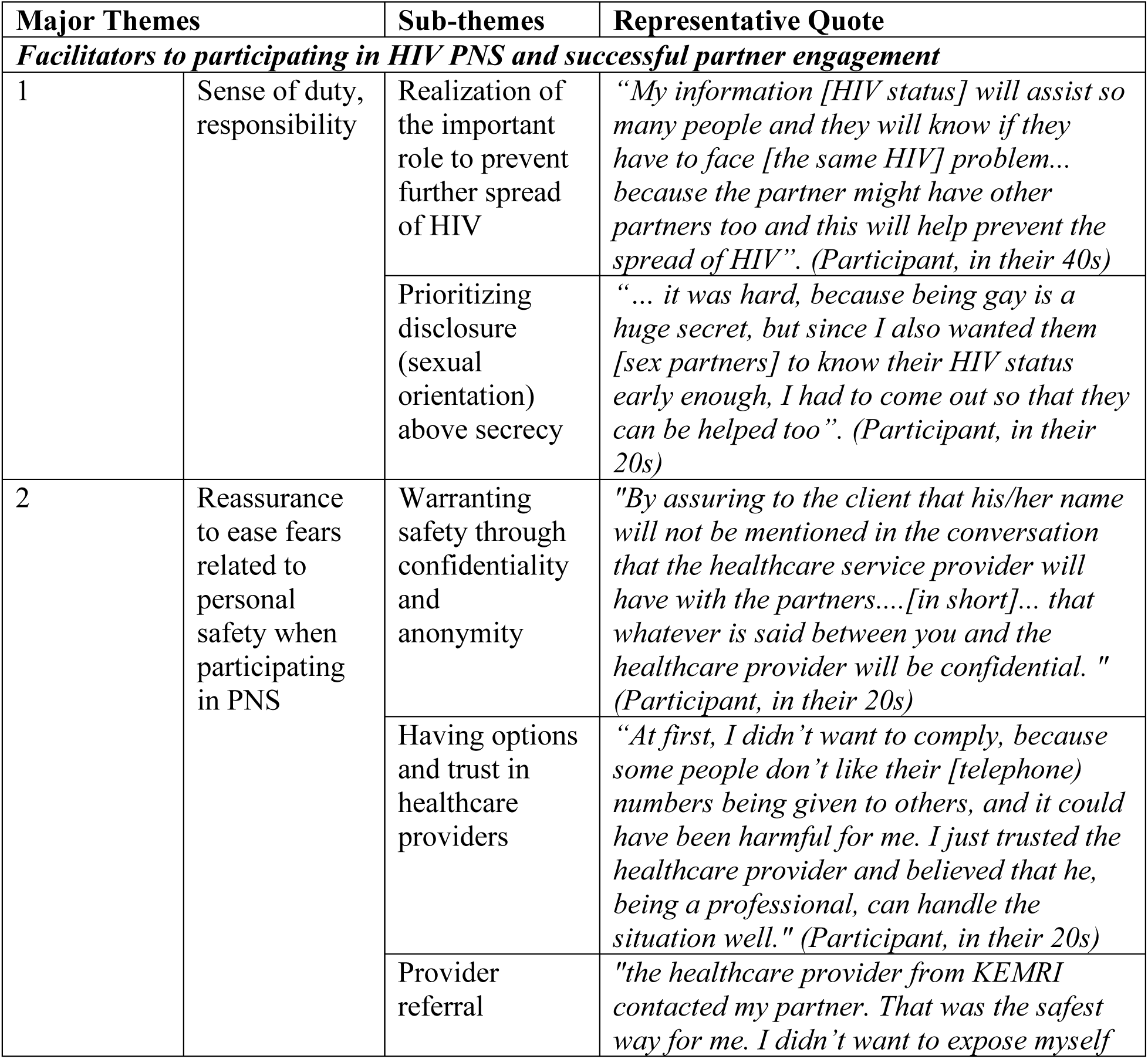
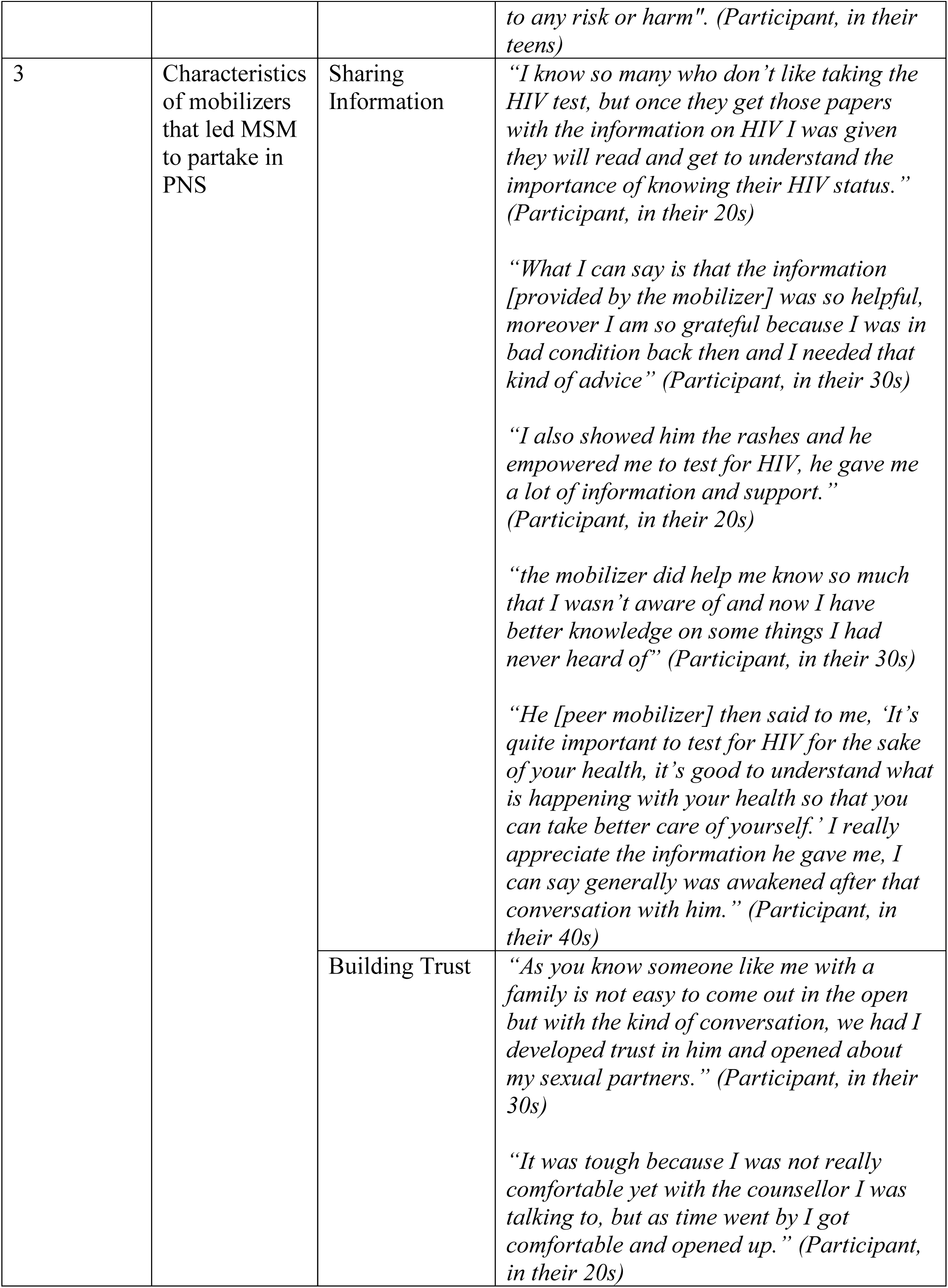
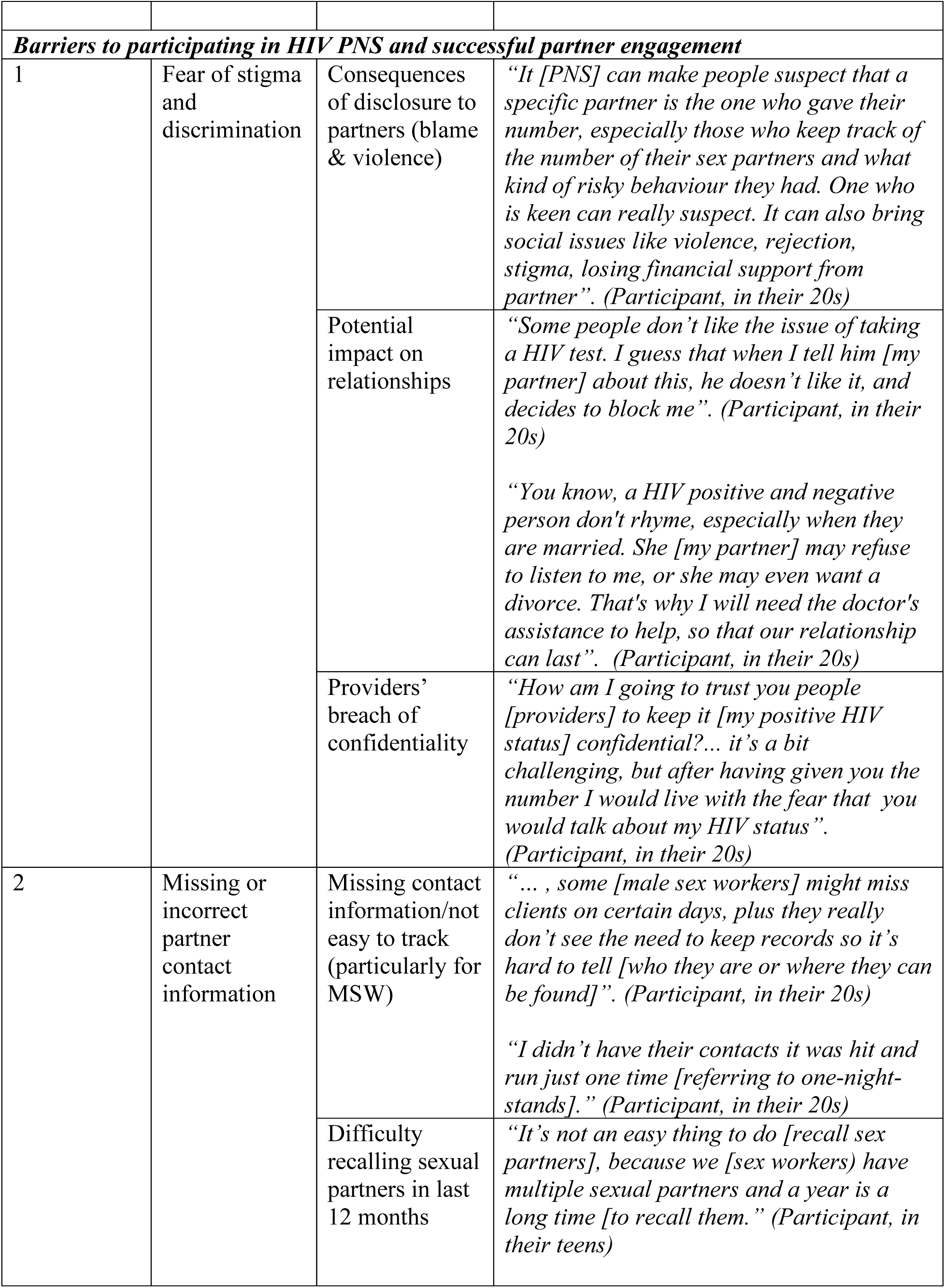
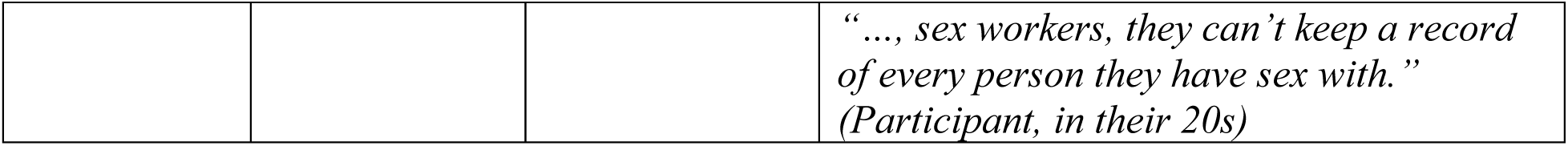
Summary of themes, sub-themes, and representative quotes identified from in-depth interview among MSM participants, coastal Kenya.

#### Barrier 1: Fear of stigma and discrimination

Participants described various concerns related to participating in PNS. Among the most notable concerns was the potential negative repercussions of disclosure to partners, which could lead to blame and violence. Despite their awareness of the confidential nature of provider-referral, index participants worried about the possibility that their partners could determine who shared their contact information to the PNS healthcare provider and, by extension, who potentially exposed them to HIV. The potential consequences associated with this indirect disclosure was described by one participant as follows:

> *“It [PNS] can make people suspect that a specific partner is the one who gave their number, especially those who keep track of the number of their sex partners and what kind of risky behaviour they had. One who is keen can really suspect. It can also bring social issues like violence, rejection, stigma, losing financial support from partner.” (Participant, in their 20s)*

Index participants who considered index referral expressed concern surrounding the potential impact the PNS program could have on their relationships with partners. Additionally, trust in healthcare providers to maintain participant confidentiality was questioned. For example, one participant noted:

> *“How am I going to trust you people [providers] to keep it [my positive HIV status] confidential?… It’s a bit challenging, but after having given you the number [of sex partners] I would live with the fear that you would talk about my HIV status.” (Participant, in their 20s)*

No participant reported experiencing any IPV or other social harms.

#### Barrier 2: Missing or incorrect partner contact information

Participants engaging in transactional sex noted the particular challenge they faced with keeping track of sexual partner contact information, and how this was not always feasible due to the volume or frequency of paid sex acts. For example, one participant stated:

> *“… some [male sex workers] might miss clients on certain days, plus they really don’t see the need to keep records, so it’s hard to tell [who they are or where they can be found].” (Participant, in their 20s)*

This was additionally challenging for one-time sexual partners, whom the index participant might not know well or might not be able to recall with accuracy. Participants also reflected on the difficulty in recalling sexual partners over the requested time frame of 12 months, which was considered a lengthy period for recalling one’s sexual episodes. This was especially true for men engaging in sex work. As one participant described:

> *“It’s not an easy thing to do [recall sex partners], because we [sex workers] have multiple sexual partners and a year is a long time [to recall them].” (Participant, in their teens)*

#### Facilitator 1: Reassurance of personal safety

Participants’ decision to partake in PNS was facilitated through reassurance by peer mobilizers or health providers to mitigate initial fears related to personal safety and possible repercussions or retaliation if their partners were able to identify them. For example, one participant noted the benefit of pre-PNS counseling provided by health care providers in the clinic:

> *"By assuring to the client that his/her name will not be mentioned in the conversation that the healthcare service provider will have with the partners….[in short]… that whatever is said between you and the healthcare provider will be confidential." (Participant, in their 20s)*

As described above, this reassurance was provided by peer mobilizers and/or health providers by explaining the confidentiality and anonymity that could be achieved through provider-referral PNS. Provider-referral PNS was recognized as a particularly important method to ensure personal safety among many participants. As one participant described:

> *"The healthcare provider from KEMRI contacted my partner. That was the safest way for me. I didn’t want to expose myself to any risk or harm." (Participant, in their teens)*

The ability to trust providers and remove themselves from challenging and possibly threatening conversations with recent sexual partners was a key facilitator for participation in provider-referral PNS among participants.

#### Facilitator 2: Characteristics of peer mobilizers that led MSM to partake in PNS

Participants reiterated the important role that peer mobilizers played in facilitating and empowering them to get tested for HIV, which then led to their participation in PNS. The educational information shared by mobilizers was described by participants as empowering, helpful, and awakening. Participants expressed their gratitude for the information and advice provided by the mobilizers which, for many, served as the push they needed to seek HIV testing. For example, one participant stated:

> *“What I can say is that the information [provided by the mobilizer] was so helpful, moreover I am so grateful because I was in bad condition back then and I needed that kind of advice.” (Participant, in their 30s)*

Additionally, the peer mobilizers’ commitment to gaining the trust of participants was described by participants as an important characteristic that promoted their involvement in HIV testing and PNS. Due to the existing burden of stigma and discrimination that MSM face, the process of opening up to the peer mobilizers was a process that took time and required patience and perseverance of the part of peer mobilizers. For example, one participant described:

> *“As you know someone like me with a family is not easy to come out in the open - but with the kind of conversation we had, I developed trust in him and opened about my sexual partners.” (Participant, in their 30s)*

Another participant noted that *“It was tough because I was not really comfortable yet with the counsellor I was talking to, but as time went by I got comfortable and opened up.” (Participant, in their 20s).* Providing time and space to enable these supportive conversations with trained peer mobilizers were therefore important characteristics to engage index participants in HIV testing and encourage them to proceed with PNS.

#### Facilitator 3: Sense of duty/responsibility to reducing transmission and facilitating partners’ testing and linkage to service

Another strong motivator for PNS described by participants was the realization of the important role they could play in preventing the further spread of HIV by participating in PNS. This sense of responsibility supported some to share contact information about their male sexual partners and reveal their sexual orientation to the peer mobilizer. For example, one participant stated:

> *“… it was hard, because being gay is a huge secret, but since I also wanted them [sex partners] to know their HIV status early enough, I had to come out so that they can be helped too….because the partner might have other partners too and this will help prevent the spread of HIV.” (Participant, in their 20s)*

## Discussion

This study provides insight into the experiences of MSM in coastal Kenya who were newly diagnosed with HIV and who utilized PNS to engage their past-year sexual partners in HIV testing. Overall, PNS was found to be a feasible and promising approach for reaching MSM in this setting with potential unrecognized exposure to HIV. Among our small cohort of 27 MSM, over 100 sexual partners were identified, of which 56% were subsequently enrolled in our study to assess their HIV status and support further PNS if needed. Of those partners enrolled in the study, 22.7% were newly diagnosed with HIV. This represents an impressive yield and points to the feasibility of utilizing PNS to identify new cases of HIV among a hard to reach, high-incidence population of MSM in coastal Kenya. Additionally, our qualitative findings revealed important facilitating factors for engaging MSM in PNS, which largely relate to the use of peer-led mobilization strategies to build trusting relationships with MSM and the importance of provider-referral PNS as a strategy. In this study, no participant reported experiencing any social or physical harm, intimate partner violence, stigma, discrimination, or negative impacts on existing relationships. Further research is necessary to explore strategies to optimize safety and minimize the potential for any risks – physical, psychological, socio-economic – related to PNS services for MSM in this setting and how to optimize effectiveness and scaling of these strategies.

Primary concerns expressed by participants related to the confidentiality of their personal identity, HIV status, and/or sexual orientation throughout the PNS process. This is consistent with the existing literature on this topic among MSM in other LMIC contexts [30, 31]. In order to overcome such barriers to participation, PNS strategies must be implemented with awareness of any possible harms to index participants and strategies relevant to local conditions that can minimize or address harms that do occur. Once these strategies are developed, MSM needed to receive continued reassurance from healthcare providers and peer mobilizers that their sensitive and deeply personal information would remain confidential. This process of trust-building was noted by participants as a key facilitating factor influencing their decision to participate in PNS and required several follow-up meetings.

The importance of provider-referral among MSM was underscored by the high frequency in selection of this strategy by index participants, regardless of type of partner. MSM reiterated the applicability of this strategy to their circumstances due to their ability to remain anonymous and protected from interpersonal harm or retaliation related to disclosure. The ability to choose from different PNS strategies, as well as opt for different strategies with different sexual partners, was expressed as an attractive feature of PNS. For example, index referral was more commonly used for reaching steady sexual partners, whereas provider referral was more commonly selected for reaching casual partners. This suggests that index participants’ level of comfort and familiarity with their partners will likely inform their choice of PNS strategy.

Notable barriers to PNS included the difficulty in recalling partner contact information over a one-year time frame and the loss or unavailability of partner contact information. These concerns were particularly pronounced among MSM who engaged in sex work but might also be relevant to other MSM who have high numbers of casual partners. Special consideration and further tailoring/adapting PNS will be needed for these MSM sub-groups. For example, shortening the recall period from 12 months to 3 months or asking MSM to share the locations where they typically meet such partners for more targeted mobilization strategies can serve as potential strategies to mitigate these challenges.

These findings contribute to the ongoing effort to fill the existing literature gap on PNS implementation with MSM in SSA. Our study findings largely corroborate the existing literature surrounding best practices for engagement with MSM in HIV testing, care, and treatment more generally. This includes the importance of maintaining confidentiality and privacy of MSM to effectively engage in HIV-related services, leveraging peers and pre-existing relationships to initiate and maintain engagement with MSM, and the fear of stigma, discrimination and harm held by MSM engaging in HIV-related services [24, 25, 26]. Leveraging a peer-led mobilization strategy to build trusting relationships with MSM participants was a particularly important facilitating factor influencing participants decision to seek HIV testing and participate in PNS. Peer-mobilizers built trusting relationships with MSM through employing patience and perseverance, sharing informative HIV-related information with participants, and leveraging existing relationships/networks. The use of peers to mobilize MSM participants in this study must be noted as a key strategy to engage MSM and their sexual partners in PNS. This finding also supports our hypothesis that peer-led HIV awareness can facilitate the uptake of PNS among MSM in this context.

This study yielded new insights to the literature on PNS and HIV prevention services for MSM in highly stigmatizing contexts. First, participants recognized that their agency in PNS plays an important in preventing HIV among other MSM in their setting, and this motivation can be a lever to help overcome initial apprehensions related to PNS services in MSM communities. All participants unanimously agreed with the importance of PNS to either provide preventative measures for HIV-negative sexual partners or ART to HIV-positive sexual partners. But the internal realization and sense of collective duty was also reiterated and seemed to overpower hesitations that some MSM had with sharing their sexual orientation with peer mobilizers/providers. The urge to prioritize others’ health over the need to preserve secrecy and conceal stigmatized identities was unexpected given prior research on the primacy of MSM identity concealment as a core motivation in this context [31]. This insight is important as peer mobilizers and healthcare providers who attempt to engage MSM to participate in PNS can leverage this sense of duty. This can be achieved by empowering MSM to recognize the agency they have in protecting the lives and well-being of others in their community. Another key finding was the need to tailor PNS among the diverse sub-groups of MSM in this context. This was particularly evident among MSM engaging in sex work, for whom it was simply not feasible or appropriate to ask them to provide or recall all of the sexual partners they had within the past year. Recognizing the diversity that exists within the broad category of ‘MSM’ will be particularly relevant as efforts to roll out PNS among MSM in Kenya progress.

Limitations to this research must be acknowledged. First, participants’ responses to both quantitative and qualitative data collection phases may have been influenced by social desirability. Second, findings might not transfer to other populations and settings in Kenya or elsewhere in SSA that lack infrastructure for conducting HIV research and programs with MSM or other stigmatized populations. Indeed, this study was built on existing relationships established between researchers, providers, peer mobilizers and MSM in the coastal Kenyan context, which enabled program delivery and evaluation. Lastly, the research protocol did not include ascertainment of information on female sexual partners of bisexual MSM; this is a critical gap and an important perspective that should be addressed in future research.

In conclusion, findings from this study further support the feasibility and promise of PNS among MSM in Kenya. Findings underscore the need for choice and personalization in PNS strategies. Although both provider-assisted and peer-led PNS strategies were commonly selected by index participants, many chose to personally contact their steady partners. Findings also highlight the need for training peer counselors and providers in principles of confidentiality and trust building in order to support index participants in the complex PNS process.

## Data Availability

All data produced in the present study are available upon reasonable request to the authors.

## Acknowledgments

The authors would like to thank those who gave their time to participate in the study and share their experience.

## Supporting Information

S1 Appendix. Intimate Partner Violence Screening Tool

S2 Appendix. Semi-structured Interview Guide

## References

1. World Health Organization. HIV data and statistics [Internet]. Geneva: WHO; 2024 [cited 2024 Aug 12]. Available from: https://www.who.int/teams/global-hiv-hepatitis-and-stis-programmes/hiv/strategic-information/hiv-data-and-statistics.

2. Sundararajan, R., Ponticiello, M., Nansera, D., Jeremiah, K., & Muyindike, W. (2022). Interventions to Increase HIV Testing Uptake in Global Settings. Current HIV/AIDS reports, 19(3), 184–193. 10.1007/s11904-022-00602-4

3. Cherutich, P., Golden, M. R., Wamuti, B., Richardson, B. A., Ásbjörnsdóttir, K. H., Otieno, F. A.,… & aPS Study Group. (2017). Assisted partner services for HIV in Kenya: a cluster randomised controlled trial. The lancet HIV, 4(2), e74–e82.

4. Brown, L. B., Miller, W. C., Kamanga, G., Nyirenda, N., Mmodzi, P., Pettifor, A.,… & Hoffman, I. F. (2011). HIV partner notification is effective and feasible in sub-Saharan Africa: opportunities for HIV treatment and prevention. Journal of acquired immune deficiency syndromes (1999), 56(5), 437.

5. Myers, R. S., Feldacker, C., Cesár, F., Paredes, Z., Augusto, G., Muluana, C.,… & Golden, M. R. (2016). Acceptability and effectiveness of assisted human immunodeficiency virus partner services in Mozambique: results from a pilot program in a public, urban clinic. Sexually transmitted diseases, 43(11), 690–695.

6. Centers for Disease Control and Prevention. Index Testing Minimum Requirements (CDC) [Internet]. Washington, D.C.: U.S. Department of State; 2024 [cited 2024 Aug 12]. Available from: https://www.state.gov/wp-content/uploads/2024/01/003.CDC-REDCap.pdf.

7. World Health Organization. WHO recommends social network-based HIV testing approaches for key populations as part of partner services package: policy brief [Internet]. Geneva: World Health Organization; 2019 [cited 2024 Aug 12]. Available from: https://iris.who.int/bitstream/handle/10665/329964/WHO-CDS-HIV-19.32-eng.pdf?sequence=1.

8. John SA, Starks TJ, Rendina HJ, Parsons JT, Grov C. High willingness to use novel HIV and bacterial sexually transmitted infection partner notification, testing, and treatment strategies among gay and bisexual men. Sex Transm Infect. 2020;96(3):173–176. doi:10.1136/sextrans-2019-053974

9. Udeagu CN, Shah S, Toussaint MM, Pickett L. Sociodemographic Differences in Clients Preferring Video-Call Over In-person Interview: A Pilot Study of HIV Tele-partner Services. AIDS Behav. 2017;21(11):3078–3086. doi:10.1007/s10461-016-1586-4

10. Edelman EJ, Cole CA, Richardson W, Boshnack N, Jenkins H, Rosenthal MS. Opportunities for improving partner notification for HIV: results from a community-based participatory research study. AIDS Behav. 2014;18(10):1888–1897. doi:10.1007/s10461-013-0692-9

11. Kerani, R., Fleming, M. & Golden, M. (2013). Acceptability and Intention to Seek Medical Care After Hypothetical Receipt of Patient-Delivered Partner Therapy or Electronic Partner Notification Postcards Among Men Who Have Sex With Men. Sexually Transmitted Diseases, 40 (2), 179–185. doi: 10.1097/OLQ.0b013e31827adc06.

12. Mimiaga MJ, Reisner SL, Tetu AM, et al. Partner notification after STD and HIV exposures and infections: knowledge, attitudes, and experiences of Massachusetts men who have sex with men. Public Health Rep. 2009;124(1):111–119. doi:10.1177/003335490912400114

13. Golden MR, Hogben M, Potterat JJ, Handsfield HH. HIV partner notification in the United States: a national survey of program coverage and outcomes. Sex Transm Dis. 2004;31(12):709–712. doi:10.1097/01.olq.0000145847.65523.43

14. Hogben, M., McNally, T., McPheeters, M., Hutchinson, A. B., & Task Force on Community Preventive Services. (2007). The effectiveness of HIV partner counseling and referral services in increasing identification of HIV-positive individuals: a systematic review. American journal of preventive medicine, 33(2), S89–S100.

15. Nguyen TTV et al. Community-led HIV testing services including HIV self-testing and assisted partner notification services in Vietnam: lessons from a pilot study in a concentrated epidemic setting. Journal of the International AIDS Society 2019, 22(S3):e25301

16. Dijkstra M, Mohamed K, Kigoro A, et al. Peer Mobilization and Human Immunodeficiency Virus (HIV) Partner Notification Services Among Gay, Bisexual, and Other Men Who Have Sex With Men and Transgender Women in Coastal Kenya Identified a High Number of Undiagnosed HIV Infections. Open Forum Infect Dis. 2021;8(6):ofab219. Published 2021 Apr 29. doi:10.1093/ofid/ofab219

17. Johnson LF, van Rensburg C, Govathson C, Meyer-Rath G. Optimal HIV testing strategies for South Africa: a model-based evaluation of population-level impact and cost-effectiveness. Sci Rep. 2019;9(1):12621. Published 2019 Sep 2. doi:10.1038/s41598-019-49109-w

18. Alam, N., Chamot, E., Vermund, S. H., Streatfield, K., & Kristensen, S. (2010). Partner notification for sexually transmitted infections in developing countries: a systematic review. BMC public health, 10(1), 1–11.

19. O’Bryan G, Chirairo H, Munyayi F, Ensminger A, Barnabee G, Dzinotyiweyi E, Mwandingi L, Ashipala L, Forster N, O’Malley G, Golden M. Assisted Partner Notification Services in Namibia: Comparison of Case-Finding in Persons With New and Previously Diagnosed Human Immunodeficiency Virus, and Success as a Platform for PrEP Referral. Sexually Transmitted Diseases. 2024 Mar 1;51(3):214–9.

20. National AIDS and STI Control Programme A guidance document for the delivery of HIV Self-Testing and assisted partner notification services in Kenya. Ministry of Health, Kenya. 2018:1–84.

21. AVAC: Global Advocacy for HIV Prevention. Activism in Action: Week 1 reporting from the frontlines of the PEPFAR planning process. January 2020. https://www.avac.org/blog/activism-action-week-1-reporting-frontlines-pepfar-planning-process

22. Shangani S, Naanyu V, Operario D, Genberg B. Stigma and healthcare-seeking practices of men who have sex with men in Western Kenya: a mixed-methods approach for scale validation. AIDS patient care and STDs. 2018 Nov 1;32(11):477–86.

23. Maleke K, Daniels J, Lane T, Struthers H, McIntyre J, Coates T. How social stigma sustains the HIV treatment gap for MSM in Mpumalanga, South Africa. Global health promotion. 2019 Dec;26(4):6–13.

24. Mbeda C, Ogendo A, Lando R, Schnabel D, Gust DA, Guo X, Akelo V, Dominguez K, Panchia R, Mbilizi Y, Chen Y. Healthcare-related stigma among men who have sex with men and transgender women in sub-Saharan Africa participating in HIV Prevention Trials Network (HPTN) 075 study. AIDS care. 2020 Aug 2;32(8):1052–60.

25. Shangani, S., Escudero, D., Kirwa, K., Harrison, A., Marshall, B., & Operario, D. (2017). Effectiveness of peer-led interventions to increase HIV testing among men who have sex with men: a systematic review and meta-analysis. AIDS care, 29(8), 1003–1013.

26. Ochonye, B., Folayan, M. O., Fatusi, A. O., Emmanuel, G., Adepoju, O., Ajidagba, B.,… & Yusuf, A. (2019). Satisfaction with use of public health and peer-led facilities for HIV prevention services by key populations in Nigeria. BMC health services research, 19(1), 1–11.

27. Yan, H., Zhang, R., Wei, C., Li, J., Xu, J., Yang, H., & McFarland, W. (2014). A peer-led, community-based rapid HIV testing intervention among untested men who have sex with men in China: an operational model for expansion of HIV testing and linkage to care. Sexually transmitted infections, 90(5), 388–393.

28. Sanders, E. J., Agutu, C., van der Elst, E., Hassan, A., Gichuru, E., Mugo, P.,… & Graham, S. M. (2021). Effect of an opt-out point-of-care HIV-1 nucleic acid testing intervention to detect acute and prevalent HIV infection in symptomatic adult outpatients and reduce HIV transmission in Kenya: a randomized controlled trial. HIV medicine.

29. Clarke V, Braun V, Hayfield N. Thematic analysis. Qualitative psychology: A practical guide to research methods. 2015 Jan 1;222(2015):248.

30. Semple SJ, Pines HA, Strathdee SA, et al. Uptake of a Partner Notification Model for HIV Among Men Who Have Sex With Men and Transgender Women in Tijuana, Mexico. AIDS Behav. 2018;22(7):2042–2055. doi:10.1007/s10461-017-1984-2

31. Yan, X., Lu, Z., Zhang, B., Li, Y., Tang, W., Zhang, L., & Jia, Z. (2020). Protecting Men Who Have Sex With Men From HIV Infection With an mHealth App for Partner Notification: Observational Study. JMIR mHealth and uHealth, 8(2), e14457. 10.2196/14457

