## Supplemental Document 1 for "Peer-assisted HIV partner notification services to strengthen index partner testing for newly diagnosed men who have sex with men in coastal Kenya"

**Assessing acceptability, feasibility, and safety of partner notification services and transmission networks of gay, bisexual, and transgender people newly diagnosed with HIV infection, a mixed-method explorative study in coastal Kenya and Nairobi**

**Topic guide for in-depth interview recently diagnosed GBT**

The objectives of the interview are to assess and explore:

- Participant’s possible experiences with PNS

- Participant’s perspectives on PNS, its impact, barriers and facilitators

**Study number : _________________________________**

**Interviewer	: _________________________________**

**Site	: _________________________________**

**Date	: _________________________________**

**Start	: _________________________________**

**End	: _________________________________**

1. **Introduction**

- Introduce yourself
- Explain goal: insight in experiences with partner notification services (PNS) and participant’s perspectives on PNS, its impact, barriers and facilitators
- Own experience, no right or wrong
- Recording? Anonymous (*With your permission, we would like to tape record the discussion for later transcription and translation to English for analysis. All information will be kept strictly confidential, and, as a participant, you will never be identified with any comment you make.)*
- Duration interview (~1 hour)
- Explain reimbursement
- Notes, ok?
- Questions?

2. **Informed consent**

- Go through informed consent form
- Questions?
- Make sure participant checks all boxes, fills first and last name, date and time, and signs
- Sign informed consent form as investigator / designee

3. **Fill out demographic form**
4. **Start audio recording**
5. **Start interview**

**Introduction**

- *We would like to learn more about partner notification services (PNS) after someone has tested positive for HIV.*
- *Partner notification means that the sex partners of someone who has tested positive for HIV are notified of their risk for HIV and are advised to test for HIV.*
  - *The person who tested positive for HIV is called the index client*
  - *PNS is a voluntary process with consent of the index client, offering the sex partners(s) voluntary HIV testing*
  - *PNS are provided by trained HCP, or trained lay persons*
  - *PNS are provided using passive or assisted approaches*
  - *During assisted PNS, a HCP notifies the partners of their risk possible exposure without revealing the identity of the index client*
- *In Kenya, it is now national policy to offer assisted PNS to someone who has tested positive for HIV. Assisted PNS have been studied in heterosexuals in Kenya and are considered safe: there was no intimate partner violence.*
- *We have little knowledge on how we could offer PNS to gay, bisexual and transgender people (GBT). In this interview, I would like to hear from you about your possible experience with PNS and whether you think PNS is feasible for GBT, and if so, how you think PNS are scalable.*

**Topic 1: HIV diagnosis experience and emotional impact**

*Focus for interviewer:* climate setting

- How are you feeling? How have you been since your diagnosis? How did you get through it? Are you comfortable talking about your experiences? Any fears / challenges?

**Topic 2: HIV testing experience and knowledge**

*Focus for interviewer on exploring knowledge and experience with HIV testing services*

- When you were diagnosed with HIV, what was the reason you tested for HIV? *(Probe for time for regular test; exposure/ transmission risk; unprotected sex; sex with someone new; starting a new relationship; suspected acute HIV infection / symptoms; other?)*
- How did you get your test results, and what was the experience like?
- Did you start treatment immediately, or later? Why?
- How did you feel about starting treatment? What can you tell me about the counselling you have received?

**Topic 3: Disclosure of HIV status to others**

*Focus for interviewer*: participant’s own experience with disclosure.

- Have you disclosed the result of your HIV test with someone? If so, with whom, and why with that particular person(s)?
- If yes, can you tell me how did you go about the disclosure? How did he/she/they react?
- If no, why haven’t you told anyone about your HIV status?
- Can I ask you whether you have disclosed to your sex partner(s)? *(Probe for steady partner, regular partner(s), casual partner(s), clients).*

**Topic 4: Sex partners and experiences with PNS**

*I would like to talk with you about your sex partner(s) you had before you were diagnosed with HIV. Most GBT have multiple sex partners, so I would like you to first think of the person you most recently had sex with before you were diagnosed with HIV. This also includes possible clients. We will then move on to the person you had sex with before this this person. I would like to ask you to share details about up till 5 sex partners with me.*

*Now I would like to ask you to think of person you had sex with most recently before you were diagnosed with HIV.*

- Can you tell me a bit more about this person? *(Probe for man or woman, type of partner: primary, casual, one-off, client)*
- Where did you meet? *(Probe for: through friends/family, social media, hotspot)*
- When did you first had sex with him/her? When was the last time?
- Do you know his/her HIV status?
- Do you think he/she knows his/her HIV status?
- Have you told or will you tell this person that you have been diagnosed with HIV?

*If no:*

- If you would tell this person, how do you think it might affect your relationship?
- If this person is not already aware that he or she is HIV-infected, do you think he/she will be willing to get tested? To start medications? *(Probe for options for contact tracing, if there are any)*

*If yes:*

- How was your partner notified of the possibility of him/her having had a risk exposure with an HIV-infected partner?
- How did he/she take it? Did he/she accept to get tested and what was the outcome?
- Was there OST provided to this partner?

If so, how do you feel about this approach?

*Now I would like to ask you to think of the person you had sex with before the person we just discussed.*

- Can you tell me a bit more about this person? *(Probe for man or woman, type of partner: primary, casual, one-off, client)*
- Where did you meet? *(Probe for: through friends/family, social media, hotspot)*
- When did you first had sex with him/her? When was the last time?
- Do you know his/her HIV status?
- Do you think he/she knows his/her HIV status?
- Have you told or will you tell this person that you have been diagnosed with HIV?

*If no:*

- If you would tell this person, how do you think it might affect your relationship?
- If this person is not already aware that he or she is HIV-infected, do you think he/she will be willing to get tested? To start medications? *(Probe for options for contact tracing, if there are any)*

*If yes:*

- How was your partner notified of the possibility of him/her having had a risk exposure with an HIV-infected partner?
- How did he/she take it? Did he/she accept to get tested and what was the outcome?
- Was there OST provided to this partner?

If so, how do you feel about this approach?

***Repeat up till five partners.***

- Have you shared with anyone else the fact that you were recently diagnosed with HIV infection?
- Have you shared your HIV status with any other sex partner(s)?
- If so, whom have you told? How much have you shared? Why did you want them to know?
- If not, whom do you want to tell? Why do you want them to know? / Are you ready to share? How much do you think they are ready to hear?
- How has / will disclosing your HIV status affect you, and how has / will it affect the people you tell? *(probe for: challenges)*

**Topic 5: Estimating the number of partners**

*Kenyan national policy recommends notification of* ***all*** *sex partners in the last 12 months before HIV was diagnosed.*

- To what extent do you think it would be possible for you to recall and notify all sex partners from the 12 months before your HIV diagnosis? Why? *(Probe for: too much time ago, difficult to remember, too many partners, anonymous partners)*
- Do you think it would be possible to give an estimation of how many sex partners you’ve had in the 12 months before your HIV diagnosis? If yes, could you give a number? If not, why not? *(Probe for: not feeling comfortable to share this information with a HCP, being afraid to be stigmatised or judged upon)*
- To what extent do you think you would be willing to share the **name and contact information** these sex partners with a HCP? *(Probe for: barriers to share, confidentiality, fear of being known as HIV positive)*

**Topic 6:** **Barriers and facilitators in offering partner notification services to GBT**

- Could you tell us of some ways that helped (or could help) you disclose / invite your partner(s) for testing? *(Probe for the perception of assisted partner notification, including barriers and facilitators)*

*As I said before, PNS means that the sex partner of someone who has tested positive for HIV are notified of their risk for HIV and are advised to test for HIV.*

*The WHO recommends PNS and recently the Kenyan MOH adopted the approach, which is now recommended for clients who test HIV positive.*

*Focus for interviewer on preferred PNS options, and why to decline PNS.*

If PNS were to be provided to GBT who test HIV positive, what are your thoughts?

- What challenges do you foresee?

*(Probe* *for* ***multiple or anonymous*** *sex partners; index clients may not remember their sex partners, or not have contact information of the sex partners)*

*(Probe for* ***reluctance*** *to share contact information with HCP;* ***fear to disclose*** *their sexuality; fear for break-up of relationship; confidentiality issues; partners may not want to test for HIV; fear for being blamed for bringing HIV into the relationship*; *other?)*

- What do you think can facilitate uptake of PNS among GBT?
- *(Probe also for emotional and social benefits)*
- Do you foresee any positive outcomes or benefits? What would these positive outcomes or benefits be? (*Probe for benefit of HIV care for positive partners, or PrEP for negative partners; other?)*

**Topic 7: Safety of partner notification services**

*Focus for interviewer on social harm or other adverse events following voluntary PNS. What does the participant think of being at risk of social harm or physical violence?*

*If partner notification not done:*

- To what extent would you be concerned about safety when PNS would be done? How come? *(Probe for the potential risk of violence/ harm; discrimination; stigma; other?)*
- Are there any other risks you would be concerned about? *(Probe for fear of violence; physical/psychological/sexual/emotional, or controlling behaviours; losing a job or a home; being rejected by family members; losing clients from sex work)*
- How do you think these risks could be averted?

*If partner notification already done:*

- Did you experience any safety issues or other harms as a result of notifying your partner(s)? *(Probe for violence; physical/psychological/sexual/emotional, or controlling behaviours; losing a job or a home; being rejected by family members; losing clients from sex work)*
- If so, what kind? What happened? When? With whom?
- Do you think this could have been averted? How?

**Topic 8: Practical aspects of PNS implementation**

*Focus for interviewer on support for effective and safe PNS.*

*As we discussed before, there are different methods to notify sex partners of a person who is diagnosed with HIV.*

- ***First****, the index client can contact his partner(s) and advise them to do an HIV test.*
- ***Second****, the index client can also provide an HIV self-test to his partner(s), and advise them to use this self-test.*
- ***Third,*** *the index client agrees to notify partners within a specified time period, and if this is not done the HCP will proceed to provider referral.*
- ***Fourth****, the index client can provide name and contact information of his partners to a HCP. The health provider will contact the partners by phone, through text message or in person and will advise the partners to do an HIV test. The name or any details of the person who was diagnosed with HIV are not disclosed to the partners.*
- What do you think about these different methods for PNS?
- Would you prefer one over another? Why?
- Based on your experiences, how best do you think PNS can be implemented for GBT?
- Could you think of another strategy for PNS? If so, what other strategy? (*Probe for social media, peers, OST, hotspots, through LGBT organisations; other?)*
- If you imagine PNS being part of standard HIV care, what would you envision a good moment to discuss PNS after someone is diagnosed with HIV? *(Probe for “on the same day” after HIV diagnosis; why / why not?)*

*(Also probe for “later”, if later, how much later after HIV diagnosis? Why?)*

- Could you come up with communication strategies for HCP to discuss PNS with a person who tested positive for HIV? *(Probe for: precise wordings on how to introduce the topic, how to discuss the number of partners and name and contact information, how to build trust on confidentiality)*
- If you were asked to give advice to designing and implementing PNS for GBT in Kenya, what would you recommend?
- Is there anything else, you would like to discuss about PNS for GBT?

*Kindly, thank the participant for sharing his views and experiences.*

*END.*

6. **Stop audio-recording**
7. **Provide reimbursement**
