## Supplemental Document 2 for "Peer-assisted HIV partner notification services to strengthen index partner testing for newly diagnosed men who have sex with men in coastal Kenya"

**Form 4: Partnership Form Partner Services Study**

*To be filled out by the research counsellor, use one form for each partnership.*

*Page 1 is confidential and will only be used during the partner notification process. After completion of partner notification services, page 1 will be destructed.*

Study number **index participant**

Study number **partner**

**A.	Contact details index participant**

Name index participant: ………………………………………………………

Address / location: ……………………………………………………………

Phone number:………………………………………………………………..

**B.	Contact details partner**

Name partner: …………………………………………………………………

Address / location:……………………………………………………………

Phone number:………………………………………………………………..

Email:……………………………………...................................................

Social media account (Grindr, Romeo, Facebook):..……………………..

Hotspot: ……………………………………...............................................

Description of physical appearance: :……………………………………...

### Part A: Sexual partner

*To be filled out by the research counsellor during the full partner notification services interview with the index participant.
Use one Partnership Form for each partner.*

1. **Gender identity of partner:**

☐	Male

☐	Female

☐	Transgender female (male to female)

☐	Other: ……………………………………

2. **Type of partner:**

☐	Steady partner (a sexual partner with whom the index participant is or was in the steady relationship)

☐	Casual partner (a sexual partner with whom the index participant had sexual intercourse more than once, but is not considered a steady partner by the index participant)

☐	One-off partner (a sexual partner with whom the index participant had sexual intercourse once)

☐	Regular client (a sexual partner with whom the index participant had sexual intercourse more than once, and has paid the index participant for sex)

☐	One-off client (a sexual partner with whom the index participant had sexual intercourse once, and has paid the index participant for sex)

3. **Date of last sexual contact of index participant and partner (day/month/year):** (oral, vaginal, or anal) ___________________________
4. **Is the partner eligible for PEP?**

☐	No	☐	Yes

5. **Has this partner physically hurt you?**

☐	Ever	☐	In past month	☐	Never

6. **Has this partner threatened, frightened, or insulted you, or treated you badly?**

☐	Ever	☐	In past month	☐	Never

7. **Has this partner forced you to participate in sexual activities that made you feel uncomfortable?**

☐	Ever	☐	In past month	☐	Never

8. **Do you think any of these things could happen to you if this partner would be advised to get tested?**
   *This could be done without revealing your identify.*

☐	No	☐	Yes

9. **Do you think any other (social) harms could happen to you if this partner would be advised to get tested?**
   *This could be done without revealing your identify.*

☐	No	☐	Yes, explain: ___________________________________

10. **What IPV risk category is this partnership in?**

☐	High (if question 4 or 5 or 6 was “in past month”)
☐	Moderate (if question 4 or 5 or 6 was “ever”)
☐	Low (if question 4 or 5 or 6 were “never”)

11. **Is this partnership eligible for partner notification services?**

☐	No (if question 10 was “high”)	☐	Yes

12. **Will the partner be notified for HIV?**

☐	No, the index participant knows the partner is HIV positive *-> discuss with the index participant if the partner would be willing to provide a blood sample for phylogenetic analysis and agree on an approach to invite the partner. Skip to Q20.*

☐	No, the partner is already notified for HIV *Skip to Q14*

☐	No, the partner recently got tested *Skip to Q14*

☐	No, the risk for IPV is high

☐	No, the index participant does not have any contact information or hotspot location of the partner

☐	No, other reason: .....................................

☐	Yes

13. **How will the partner be notified?**

*Depending on the preference of the index participant.*

☐ A KEMRI counsellor will contact the partner and invite him/her for HIV testing, without revealing the identity of the index participant; -> *collect as much contact information of the partner needed to notify the partner (page 1)*

☐ A peer mobiliser will offer OST to the partner, without revealing the identity of the index participant; -> *collect as much contact information of the partner needed to notify the partner (page 1)*

☐ A peer mobiliser will offer OST at the hotspot of the partner, including to the partner, without revealing the potential risk exposure of the partner, and without revealing the identity of the index participant; -> *collect as much contact information of the partner needed to notify the partner (page 1)*

☐ A KEMRI counsellor will support the index participant to invite the partner for testing / to come for couples counselling; -> *issue numbered card for partner with invite for HTC and agree on a date by which the index participant has notified the partner* **(day/month/year):** ___________________________

☐ A peer mobiliser will support the index participant to invite the partner for testing / to come for couples counseling; -> *issue numbered card for partner with invite for HTC and agree on a date by which the index participant has notified the partner* **(day/month/year):** ___________________________

☐ The index participant will provide OST to the partner and advise the partner to seek confirmatory testing, with optional disclosure; -> *issue OST Tracking Form for partner and agree on a date by which the index participant has provided OST to the partner* **(day/month/year):** ___________________________

**In case the index participant was also diagnosed with an STI:**

☐ A KEMRI counsellor will contact the partner and inform the partner of his/her potential risk exposure of an STI and invite him/her for STI/HIV testing, without revealing the identity and HIV status of the index participant; -> *collect as much contact information of the partner needed to notify the partner (page 1)*

☐ The index participant will advise the partner to seek STI testing and treatment at KEMRI, where the partner will also be offered HIV testing. -> *issue numbered card for partner with invite for HTC and agree on a date by which the index participant has notified the partner* **(day/month/year):** ___________________________

☐ Other: …………………………………………….

Comments: _____________________________________________________________________________

### Part B: Evaluation

*To be filled out by the research counsellor after completion of partner notification services.*

14. **Who notified the partner for HIV?**

☐	KEMRI health provider (HIV testing)

☐	Peer mobiliser offered OST to the partner

☐	Peer mobiliser provided OST at the hotspot of the partner, including to the partner

☐	Index participant with support of a KEMRI health provider

☐	Index participant with support of a peer mobiliser

☐	Index participant by providing OST to the partner

☐	KEMRI health provider (STI exposure)

☐	Index participant (STI exposure)

☐	Other: …………………………………………….

15. **How was the partner notified for HIV?**

☐	Personal (face-to-face)

☐	Letter

☐	Phone call

☐	Text message

☐	Through social media

☐	Through hotspot

☐	Email

☐	Other: …………………………………………….

16. **Did the partner test for HIV?**

☐	No, the partner stated to be HIV positive – skip to question 22

☐	No, the partner did not want to test for HIV – end of questionnaire

☐	No, other reason: ……………………………… – end of questionnaire

☐	Yes – skip to question 17

☐	Unknown – end of questionnaire

17. **Where did the partner test for HIV?**

☐	KEMRI Mtwapa – skip to question 19

☐	KEMRI Malindi – skip to question 19

☐	OST – skip to question 18

☐	Unknown – skip to question 20

☐	Other: ……………………………………………. – skip to question 20

18. **Did the partner present for confirmatory testing at KEMRI after using OST?**

☐	No, skip to Q20	☐ Yes	☐ Unknown, skip to Q20

19. **HTC number of partner:** …………………………………………….
20. **What was the HIV test result of the partner?**

☐	Newly diagnosed with acute HIV infection

☐	Newly diagnosed with established (chronic) HIV infection, skip to Q22

☐	Previously diagnosed with HIV infection, skip to Q22

☐	HIV negative, skip to Q22

☐	Unknown, skip to Q22

21. **Diagnosis of acute HIV infection based on:**

☐ Negative HIV test in the previous 3 months

☐ HIV-RNA positive, no HIV antibodies

☐ HIV-RNA positive, discrepant rapid test results

22. **The HIV test results:**

☐	Were reported by the index participant

☐	Were reported by the partner

☐	Were reported by peer mobiliser

☐	Were retrieved from the mobilisation participant form or partner’s file at KEMRI

☐	Other: …………………………………………….

23. **In case the partner was eligible for PEP: did the partner start PEP?**

☐	No, explain…………………………………………….

☐	Yes, date PEP was initiated (day/month/year) …………………………………………….
